# PREVALENCE AND FACTORS ASSOCIATED WITH MATERNAL AND FETAL OUTCOMES IN WOMEN UNDERGOING CESAREAN SECTION AT THE MONO-COUFFO REGIONAL HOSPITAL CENTER BETWEEN 2020 AND 2021

**DOI:** 10.64898/2026.01.27.26344997

**Authors:** Edayé Beaudouin Jean-de-Dieu, Klikpezo Roger, Olowo Ingrid, Obossou Awade Afoukou Achille, Agonnoudé Togbédji Maurice, Amouzoun Sabine, Ezinmegnon Mahugnon Boris Philippe

**Author notes:** Corresponding author: Edaye BDJ.

## Abstract

**Introduction:** This study evaluates the maternal and fetal outcomes of cesarean sections and identifies the associated factors in a reference hospital center in Benin.

**Study Method:** This is a retrospective analytical study conducted on the medical records of women who underwent cesarean delivery at the Mono-Couffo Regional Hospital Center between 2020 and 2021. Maternal and fetal outcomes were categorized as favorable or unfavorable based on standardized criteria. Sociodemographic, obstetric, and medical data of the patients were statistically analyzed, with a significance threshold set at p < 0.05.

**Results:** The average age was 27.04 years, and 78.33% resided in rural areas. The overall cesarean section prevalence was 46.26%, with 76.78% being emergency cesareans and 23.22% scheduled. Unfavorable outcomes affected 10.84% of mothers and 17.96% of fetuses. Multiple pregnancies, urinary tract infections, a history of spontaneous abortion, and prenatal care were significantly associated with unfavorable maternal outcomes, while primiparity and nulliparity increased the risk of unfavorable fetal outcomes. These results highlight the importance of prenatal care.

**Conclusion:** Strengthening prenatal care and targeting at-risk patients would improve maternal and fetal prognoses following cesarean section.

## INTRODUCTION

Caesarean section is a surgical procedure that allows the extraction of the fetus and its appendages through a surgical opening of the uterus via a trans-abdominal approach, and rarely via the vaginal route [1]. According to the World Health Organization (WHO), the optimal caesarean section rate should range between 5% and 15% [2,3]. However, due to variations in the realities and capacities of health facilities, some degree of variation may be acceptable, but rates should not exceed 25%. Globally, the caesarean section rate among nulliparous women increased from 10% in 2001 to 16% in 2007 [1].

The ideal caesarean section rate that ensures the best maternal and neonatal benefit–risk ratio is not clearly defined in the literature [4]. It has been suggested that caesarean section rates between 5% and 10% are associated with the most favorable benefit–risk balance, whereas rates above 15% are associated with maternal and fetal risks that outweigh the expected benefits [5,6]. Population-based rates below 3% indicate underutilization of health services and suggest that some women who require caesarean delivery do not have access to it [7]. Countries with the highest maternal mortality ratios worldwide are also those with caesarean section rates below 3%. Several experiences in West Africa during the 1990s demonstrated a reduction in maternal mortality when access to caesarean section improved [8].

Some studies conducted in Africa have shown that caesarean section is associated with increased severe maternal and neonatal morbidity compared with vaginal delivery [9]. Caesarean section is a beneficial medical intervention when offered to women at high obstetric risk (Robson groups 5 to 10). Conversely, when excessively used in low-risk obstetric populations, the risks associated with the surgical procedure outweigh the expected benefits. Adverse effects are more pronounced in intrapartum caesarean sections compared with pre-labor caesarean sections [9]. The most frequent complications of caesarean section are postpartum hemorrhage and postpartum infection. Labor dystocia, which leads to prolonged labor and is a frequent indication for intrapartum caesarean section, is a known risk factor for postpartum hemorrhage. In a large North American prospective study, the rate of postpartum hemorrhage due to uterine atony was 8.3% following caesarean section [10], which was significantly higher than rates observed after vaginal delivery [11]. In contrast, pre-labor caesarean section was associated with a two-fold lower risk than intrapartum caesarean section [12], and even a lower risk than that observed after spontaneous vaginal delivery following labor [12].

Caesarean section also combines the risks of postoperative infectious morbidity inherent to any surgical procedure with those related to the specific circumstances of labor. Among primiparous women, Allen et al. showed that the risk of postpartum infection was 15 times higher compared with spontaneous vaginal delivery without instrumental assistance and 8 times higher compared with instrumental vaginal delivery [12]. Two other studies reported that rehospitalizations due to postpartum infectious complications were more frequent after caesarean delivery than after vaginal delivery, with or without instrumental assistance [13]. Beyond maternal risks, caesarean section is also associated with an increased risk of perinatal mortality related to prolonged labor, asphyxia, and sepsis [14].

It therefore appears clear that caesarean section influences maternal and fetal outcomes after childbirth. In referral hospitals, interpretation of caesarean section rates is challenging due to the role of these facilities within the national health system hierarchy. In this context, we focused on maternal and neonatal prognosis following caesarean section. The objective of this study was to assess maternal and fetal outcomes after caesarean section and to identify factors influencing maternal and neonatal outcomes following caesarean delivery at the Departmental Hospital Centers of Couffo and Mono in 2022.

## STUDY METHODS

This was a descriptive and analytical observational study with retrospective data collection covering the period from January 1, 2020, to December 31, 2021, corresponding to a two-year duration. The medical records were reviewed and the data were extracted from February 15 to February 22, 2022. The study focused on the medical records of women who delivered by caesarean section at the maternity unit of the Departmental Hospital Center (DHC) of Lokossa during this period. A simple random probabilistic sampling method was used, and the sample size was calculated using Schwartz’s formula.

The dependent variables were maternal outcome and fetal outcome among women who underwent caesarean section, coded and classified as binary variables (“Yes” or “No”). An unfavorable maternal outcome was defined as any situation in which the patient died as a result of the caesarean section, required blood transfusion, postoperative resuscitation, or emergency referral due to complications related to the procedure. An unfavorable fetal outcome was defined as any case of postoperative fetal death (excluding intrauterine fetal death), any neonatal complication, fetal distress, an APGAR score below 7, neonatal resuscitation, or neonatal referral due to complications associated with caesarean section.

The independent variables included sociodemographic characteristics (age, occupation, educational level, etc.), gyneco-obstetric data (obstetric history, indications for caesarean section, etc.), as well as maternal behavioral and individual characteristics. Data collection was carried out through a review of the medical records of patients meeting the inclusion criteria. After preliminary screening and classification of the selected records, they were sequentially numbered prior to analysis. Data were recorded using structured data collection forms and an abstraction grid, then entered into a pre-designed database using EpiData software (version 3.2, French version). Prior to final data entry, consistency checks and duplicate verification were performed.

Statistical analyses were conducted using MedCalc software (version 19.4.1, Mariakerke, Belgium) and Epi Info (version 7). Prevalence Ratios (PRs) with 95% confidence intervals (95% CIs) were used to assess associations between qualitative variables. Statistical significance was set at a p-value < 0.05. All collected data were treated with strict confidentiality and in full respect of patients’ rights. The information collected was used exclusively for the purposes of this study, in accordance with Law No. 2009-09 of May 22, 2009, on the protection of personal data in the Republic of Benin.

## RESULTS

### Frequency of Caesarean Section at the Mono Departmental Hospital Center (2020–2021)

Between 2020 and 2021 at the Mono Departmental Hospital Center (DHC), the maternity unit recorded a total of 4,442 deliveries, including 2,055 caesarean sections, corresponding to a caesarean section rate of 46.26%.

The prevalence of emergency caesarean sections was estimated at 76.78%, while scheduled caesarean sections accounted for 23.22%. These results are presented in Table I.

**Table I.** Prevalence of caesarean section types among women undergoing caesarean delivery at Mono DHC, 2020–2021.

| Type of caesarean section | Prévalence (%) | 95% CI |
| --- | --- | --- |
| Emergency | **76.78** | **[71.88 ; 81.05]** |
| Scheduled | 23.22 | [18.95 ; 28.12] |

### Age and Occupation

The mean age of women whose medical records were included in the study was 27.04 ± 5.98 years, ranging from 16 to 43 years. The most represented age group was 20–29 years, accounting for 59.13% of the sample. At the end of the study, women classified as artisans or traders represented 50.46% of the study population (Table II).

**Table II.** Distribution of women who delivered by caesarean section according to age and socio-professional category (Mono–Couffo DHC; 2020–2021)

|  | Number | Proportion (%) |
| --- | --- | --- |
| **Age (years)** |  |  |
| <20 | 25 | 07.74 |
| [20 - 30[ | 191 | 59.13 |
| [30 - 40[ | 93 | 28.79 |
| ≥40 | 14 | 04.34 |
| **Socio-professional category** |  |  |
| Farmer | 04 | 01.24 |
| Artisan/Trader | 163 | 50.46 |
| Intellectual profession | 31 | 09.60 |
| Intermediate profession | 44 | 13.62 |
| Employee | 22 | 06.81 |
| Manual worker | 06 | 01.86 |
| Unemployed | 53 | 16.41 |

### Educational Level, Marital Status, and Place of Residence

Primary education was the most frequent level of schooling, accounting for 73.07% of the women. Nearly all women were married (98.76%). The results also showed that 78.33% of the women lived in rural areas (Table III).

**Table III.** Distribution of women who delivered by caesarean section according to educational level, marital status, and place of residence (Mono–Couffo DHC; 2020–2021)

|  | **Number** | **Proportion (%)** |
| --- | --- | --- |
| **Educational level** |  |  |
| Primary | **236** | **73.07** |
| Secondary | 67 | 20.74 |
| University | 20 | 06.19 |
| **Marital status** |  |  |
| Single | 04 | 01.24 |
| Married | **319** | **98.76** |
| **Place of residence** |  |  |
| Rural | **253** | **78.33** |
| Urban | 70 | 21.67 |

### Gravidity and Parity

Table IV shows the distribution of women according to gravidity and parity. Overall, 76.78% of women were multigravida and 42.41% were multiparous.

**Table IV.** Distribution of women who delivered by caesarean section according to gravidity and parity (Mono–Couffo DHC; 2020–2021)

|  | Effectif | Proportions (%) |
| --- | --- | --- |
| **Gravidity** |  |  |
| Primigravida | 75 | 23.22 |
| Multigravida | 248 | 76.78 |
| **Parity** |  |  |
| Nulliparous | 91 | 28.17 |
| Primiparous | 70 | 21.67 |
| Multiparous | **137** | 42.41 |
| Grand multiparous | 25 | 07.74 |

### Assessment of Maternal and Fetal Outcomes

According to the predefined assessment criteria, slightly more than one in ten women (10.84%) experienced an unfavorable maternal outcome following caesarean delivery. In addition, fetal outcomes were unfavorable in 17.96% of cases (Table V).

**Table V.** Distribution of women who delivered by caesarean section according to maternal and fetal outcomes (Mono–Couffo DHC; 2020–2021)

|  | Effectif | Proportions (%) |
| --- | --- | --- |
| **Maternal outcome** |  |  |
| Unfavorable | 35 | 10.84 |
| Favorable | 299 | 89.84 |
| **Fetal outcome** |  |  |
| Unfavorable | 58 | 17.96 |
| Favorable | 265 | 82.04 |

### Factors Associated with Maternal Outcomes among Women Undergoing Caesarean Section

In this study, maternal sociodemographic characteristics were not associated with maternal outcomes following caesarean section. Factors significantly associated with unfavorable maternal outcomes included multiple pregnancy (PR = 6.41; 95% CI [2.13–19.29]; p = 0.0009), a history of spontaneous abortion (PR = 2.92; 95% CI [1.32–6.44]; p = 0.0077), and urinary tract infection (PR = 4.69; 95% CI [2.10–10.46]; p = 0.0002). Inadequate antenatal care follow-up was also associated with an unfavorable maternal outcome (PR = 2.13; 95% CI [1.04–4.35]; p = 0.0384).

**Tableau I :** Gynecological and obstetric factors according to maternal outcome among women who underwent cesarean section at the Departmental University Hospital of Mono-Couffo (CHU Lokossa; 2020– 2021)

|  | N | Maternal outcome |  | PR | 95% CI | P-value |
| --- | --- | --- | --- | --- | --- | --- |
|  |  | Unfavorable | Favorable |  |  |  |
| Type of pregnancy |  |  |  |  |  |  |
| Multiple | 15 | 6(40.00) | 09(60.00) | 6.41 | 2.13 – 19.29 | 0.0009 |
| Singleton | 308 | 29(9.42) | 279(90.58) | 1 | - | - |
| Urinary tract infection |  |  |  |  |  |  |
| Yes | 32 | 12(37.50) | 20(62.50) | 4.69 | 2.10 – 10.46 | 0.0002 |
| No | 291 | 33(11.34) | 258(88.66) | 1 | - | - |
| History of spontaneous abortion |  |  |  |  |  |  |
| Yes | 50 | 11(22.00) | 39(78.00) | 2.92 | 1.32 – 6.44 | 0.0077 |
| No | 273 | 24(08.79) | 249(91.21) | 1 | - | - |
| CPN suivi |  |  |  |  |  |  |
| Yes<sup>1</sup> | 140 | 21(15.00) | 119(85.00) | 2.13 | 1.04 – 4.35 | 0.0384 |
| No | 183 | 14(07.65) | 169(92.35) | 1 | - | - |

**Tableau VII.** Gynecological and obstetric factors according to fetal outcome among newborns of mothers who underwent cesarean section at the Departmental University Hospital of Mono-Couffo (CHU Lokossa; 2020–2021)

|  | N | Fetal outcome |  | PR | 95% CI | P-value |
| --- | --- | --- | --- | --- | --- | --- |
|  |  | Unfavorable | Favorable |  |  |  |

<sup>1</sup> Antenatal care (ANC): Yes if the mother attended at least four ANC visits

|  |  |  |  |  |  |  |
| --- | --- | --- | --- | --- | --- | --- |
| **Gravidity** |  |  |  |  |  |  |
| Primigravida | 75 | 23(30,67) | 52(69,33) | 2,17 | 1,37 – 3,43 | 0,0009 |
| Multigravida | 248 | 35(14,11) | 213(85,89) | 1 | - | - |
| **Parity** |  |  |  |  |  |  |
| Nulliparous | 91 | 26(28,57) | 65(71,43) | 4,00 | 1,61 – 9,88 | 0,0027 |
| Multiparous | 137 | 22(16,06) | 115(83,94) | 2,24 | 0,88 – 5,68 | 0,0869 |
| Grand multiparity | 25 | 05(20,00) | 20(80,00) | 2,80 | 0,88 – 8,86 | 0,0799 |
| Primiparity | 70 | 05(07,14) | 65(92,86) | 1 | - | - |
| **ATCD de césarienne** |  |  |  |  |  |  |
| Yes | 118 | 07(05,93) | 111(94,07) | 0,23 | 0,11 – 0,50 | 0,0002 |
| No | 205 | 51(24,88) | 154(75,12) | 1 | - | - |
| **History of spontaneous abortion** |  |  |  |  |  |  |
| Yes | 50 | 40(80,00) | 10(20,00) | 0,21 | 0,16 – 0,29 | <0,0001 |
| No | 273 | 48(17,58) | 225(82,42) | 1 | - | - |

## DISCUSSION

### Overall Caesarean Section Rate

The in-hospital caesarean section rate observed at the Mono–Couffo Departmental Hospital Center (46.26%) was markedly higher than the 15% threshold recommended by the World Health Organization (WHO). A study conducted by Mongbo V. [15] and published in 2016 on the quality of caesarean sections in 12 hospitals in Benin reported an average caesarean section rate of 37.6%. The high prevalence observed in our study may be explained by the role of this hospital as a referral center for complex obstetric cases in the Mono and Couffo departments and surrounding areas.

Geographical and institutional variations are known to influence caesarean section rates, particularly in settings where access to antenatal care is limited, leading to delayed management of obstetric complications. Although a high caesarean section rate may reflect improved access to emergency obstetric care, it becomes concerning when it indicates potential overuse of the procedure in cases that could be safely managed by vaginal delivery. Long-term implications include an increased risk of complications in subsequent pregnancies, such as uterine rupture and pelvic adhesions. These findings highlight the need to critically evaluate clinical practices in order to identify strategies that balance the benefits and risks of caesarean delivery, particularly through clinician training and the implementation of evidence-based protocols.

Similarly, Mylonas and Friese [16] reported a caesarean section rate of 31.7% in Germany in 2015, largely influenced by maternal request and defensive obstetric practices. In China, Deng et al. [17] reported a prevalence of 36% in 2021, with a substantial proportion of caesarean sections performed at maternal request (8.42%). Conversely, Sharma and Dhakal [18] reported a lower prevalence of 25.8% in Nepal, where indications were predominantly medical. These disparities reflect differences in health system organization, perceptions of obstetric risk, and resource availability. In our context, the referral role of the hospital likely accounts for the elevated prevalence, underscoring the need to review indications in order to reduce non-essential caesarean sections.

### Emergency versus Elective Caesarean Sections

The high prevalence of emergency caesarean sections (76.78%) compared with elective procedures (23.22%) indicates a predominantly reactive management of obstetric complications. Emergency caesarean sections are commonly associated with maternal or fetal distress, including labor dystocia, abnormal fetal heart rate patterns, and hemorrhage. Although often life-saving, these procedures carry a higher risk of maternal and neonatal morbidity and mortality compared with planned caesarean sections.

Elective caesarean sections allow for anticipation and mitigation of risks, particularly in cases of multiple pregnancies, previous caesarean delivery, or placenta previa. The observed imbalance between emergency and elective procedures highlights the need to optimize antenatal care, particularly through early identification of high-risk pregnancies to facilitate timely planning of delivery. From a scientific perspective, a key challenge lies in reducing emergency caesarean sections while maintaining effective management of obstetric complications.

Consistent with our findings, Alemu et al. [19] reported in Ethiopia in 2023 that emergency caesarean sections were associated with complications in 44% of cases. In Italy, Bevilacqua et al. [20] observed a high prevalence (85%) of elective caesarean sections among twin pregnancies, reflecting improved anticipation of obstetric risks. In contrast, Esercan et al. [21] in Turkey recommended scheduling caesarean sections after 39 weeks of gestation to reduce complications associated with emergency procedures. The predominance of emergency caesarean sections in our study may therefore reflect delays in the recognition of obstetric complications, reinforcing the need for strengthened antenatal surveillance and improved delivery planning.

### Maternal Outcomes

Unfavorable maternal outcomes were observed in 10.84% of women, reflecting significant postoperative complications such as hemorrhage, infection, or the need for postoperative resuscitation. These findings highlight the challenges faced by health systems in managing perinatal complications. Reducing such outcomes requires optimization of surgical and postoperative protocols, including effective infection prophylaxis and close monitoring of high-risk patients.

When compared with other studies, this rate falls within the range reported by referral centers in sub-Saharan Africa, although it remains a cause for concern. Alemu et al. [19] reported a substantially higher rate of 44.04% in Ethiopia, largely attributed to adverse socioeconomic conditions and limited access to quality care. In Germany, Mascarello et al. [22] found that caesarean section was associated with a significantly increased risk of maternal mortality (OR = 3.10) and postoperative infection (OR = 2.83). Conversely, Maayan-Metzger et al. [23] demonstrated that severe complications were rare in settings where well-established surgical protocols were in place. These findings emphasize the critical importance of infection prevention measures and postoperative care. In our context, the relatively lower rate of unfavorable maternal outcomes may be explained by improved surgical management within a referral hospital setting.

### Fetal Outcomes

Unfavorable neonatal outcomes were observed in 17.96% of cases, highlighting challenges related to perinatal asphyxia, prematurity, and neonatal infections. These complications may be associated with delayed interventions, emergency caesarean sections, or inadequate neonatal care. The findings underscore the need to strengthen neonatal resuscitation capacities and intensive care services in referral hospitals.

In line with our results, Keag et al. [24] reported in their 2018 meta-analysis that caesarean section was associated with an increased risk of neonatal respiratory distress (OR = 1.21). In Spain, Oros Ruiz et al. [25] found that gestational diabetes increased both caesarean section rates and the risk of neonatal asphyxia (OR = 1.5). In China, Wang et al. [26] reported a neonatal asphyxia rate of 10.2% among pregnancies complicated by congenital heart obstruction, a figure comparable to that observed in our study.

Overall, these data confirm that while caesarean section may reduce certain obstetric risks, it can also adversely affect neonatal outcomes, particularly through respiratory immaturity or preterm birth. Improved management of indications and timely decision-making may help mitigate these complications.

### Factors Associated with Maternal Outcomes

#### Multiple Pregnancies (Risk Factor)

Multiple pregnancies substantially increase the risk of maternal complications, largely due to increased mechanical and physiological stress on the uterus, placenta, and pelvic structures. Such pregnancies are frequently associated with higher rates of postpartum hemorrhage, uterine rupture, and pre-eclampsia. This association highlights the need for rigorous antenatal follow-up, including regular ultrasonographic assessment and the implementation of specific delivery protocols to anticipate potential complications. From a scientific perspective, these findings support further investigation into the benefits of planned hospitalization of term multiple pregnancies to reduce adverse maternal outcomes.

Bevilacqua et al. [20] reported in 2024 that twin pregnancies were associated with increased risks of neonatal distress and admission to intensive care units, particularly in cases of non-cephalic presentation. Earlier work by Mabie and Lavery [27] in Germany also emphasized that multiple pregnancies predispose women to complications such as placenta previa, often necessitating caesarean delivery. Collectively, these findings reinforce the importance of enhanced surveillance and anticipatory management to optimize maternal and fetal outcomes in multiple pregnancies.

#### Urinary Tract Infections (Risk Factor)

Urinary tract infections (UTIs) significantly increase the risk of adverse maternal outcomes, reflecting an increased susceptibility to postoperative infectious complications in the presence of pre-existing bacterial contamination. UTIs, which are often underdiagnosed during pregnancy, may trigger systemic inflammatory responses that complicate postoperative recovery. Prevention of unfavorable outcomes relies on systematic screening for UTIs beginning in the first trimester and prompt, effective treatment prior to delivery. Scientifically, these findings underscore the importance of integrating advanced microbiological approaches to detect and manage asymptomatic infections in high-risk pregnant women.

Mascarello et al. [22] similarly observed that infections significantly increased the risk of postoperative sepsis (OR = 2.83). In Ethiopia, Alemu et al. [19] reported higher complication rates among women living in rural areas, where infectious diseases are more prevalent and access to care is limited. These findings emphasize the critical role of systematic screening and early treatment of UTIs during pregnancy, particularly in resource-limited settings.

#### History of Spontaneous Abortion (Risk Factor)

In our study, a history of spontaneous abortion was significantly associated with an increased risk of unfavorable maternal outcomes, with a prevalence ratio (PR) of 2.92. This indicates that women with one or more previous spontaneous abortions were nearly three times more likely to experience maternal complications compared with women without such a history.

Spontaneous abortions, particularly when inadequately managed, may result in anatomical abnormalities such as intrauterine adhesions (Asherman syndrome) or uterine scarring, thereby increasing the risk of postpartum hemorrhage, uterine rupture, or infection. Furthermore, underlying causes of prior spontaneous abortions—such as hormonal disorders, chronic conditions (e.g., hypertension or diabetes), or infections—may persist and adversely affect subsequent pregnancies. Psychological stress related to prior pregnancy loss may also indirectly contribute to adverse outcomes through physiological stress pathways, including elevated cortisol levels.

These findings highlight the importance of proactive management of women with a history of spontaneous abortion, including systematic screening for uterine abnormalities and infections, as well as intensified antenatal follow-up. Alemu et al. [19] reported similar findings in Ethiopia, with significantly increased risks of postoperative maternal complications, particularly hemorrhage (OR = 3.54), among women with complicated obstetric histories. Mascarello et al. [22] also reported increased risks of postpartum infection and hemorrhagic complications associated with previous abortions. Keag et al. [24] attributed these complications to intrauterine scarring and placental disorders, such as placenta accreta, which are more frequent among women with a history of abortion. In contrast, Sharma and Dhakal [18] did not observe a significant association in Nepal, possibly due to smaller sample sizes or differences in care protocols.

### Factors Associated with Fetal Outcomes

#### Primigravidity (Risk Factor)

Primigravidity is a key risk factor for unfavorable fetal outcomes, often due to less adaptive obstetric physiology during labor. Primigravid women are at increased risk of labor dystocia and fetal distress, necessitating heightened intrapartum surveillance. Alemu et al. [19] reported that primigravidae had a significantly increased risk of neonatal complications, including prematurity and admission to neonatal intensive care units (adjusted OR = 3.476). Similarly, Keag et al. [24] observed higher rates of neonatal respiratory distress and low birth weight among primigravidae following caesarean delivery. Sharma and Dhakal [18] also reported increased frequencies of neonatal asphyxia and prolonged labor among primigravid women, suggesting the need for intensified intrapartum support. These findings are consistent with the literature identifying primigravidae as a high-risk group and support the implementation of targeted strategies, including early assessment of labor progression and enhanced clinical monitoring.

#### Nulliparity (Risk Factor)

Nulliparity is associated with increased fetal vulnerability, likely related to altered obstetric mechanics and a higher likelihood of emergency caesarean section. Mylonas and Friese [16] reported that nulliparous women were more likely to undergo emergency caesarean delivery, thereby increasing the risk of fetal complications. Bevilacqua et al. [20] also observed higher fetal morbidity rates among nulliparous women, particularly in multiple pregnancies, potentially due to limited obstetric experience and suboptimal antenatal follow-up. Mascarello et al. [22] further identified an association between nulliparity and increased intrapartum hemorrhage, which may compromise fetal oxygenation. These findings align with existing evidence and underscore the need for enhanced fetal surveillance and rapid intervention strategies when managing nulliparous women.

#### Previous Caesarean Section (Protective Factor)

The protective effect of a previous caesarean section on fetal outcomes observed in this study may be explained by more rigorous pregnancy monitoring and planned delivery strategies. Ali and Chandraharan [28] reported that well-managed pregnancies following a previous caesarean section were associated with reduced risks of fetal distress due to scheduled deliveries (OR = 0.23). In contrast, Keag et al. [24] noted a slightly increased risk of prematurity and neonatal respiratory distress, largely attributable to early planned deliveries. Deng et al. [17] reported that prior caesarean sections reduced intrapartum complications by limiting prolonged labor, although outcomes were highly dependent on obstetric protocols.

The protective effect observed in our study contrasts with findings by Keag et al. [24], possibly reflecting differences in surgical and obstetric protocols or the exclusion of severe complications (e.g., placenta previa or accreta) that may bias outcomes in other studies. These findings emphasize the importance of proactive management and intensive follow-up to improve perinatal outcomes among women with a history of caesarean section.

#### History of Spontaneous Abortion

Although a history of spontaneous abortion is often considered a risk factor, the protective association observed in our study may be explained by several hypotheses. First, women with prior spontaneous abortions often benefit from intensified medical follow-up, including frequent antenatal visits, infection screening, and early intervention in the event of complications. Second, spontaneous abortion may act as a biological selection mechanism by eliminating nonviable fetuses with severe anomalies, thereby increasing the likelihood of subsequent viable pregnancies. Third, women with prior pregnancy loss may adopt more protective health behaviors, such as improved adherence to antenatal care, healthier nutrition, and avoidance of harmful substances.

Identification of women with a history of spontaneous abortion may therefore facilitate the implementation of intensified follow-up strategies that contribute to improved fetal outcomes. Nevertheless, residual confounding by factors such as maternal age, socioeconomic status, or access to care cannot be excluded.

This finding contrasts with several studies reporting increased fetal risk among women with a history of spontaneous abortion. Sharma and Dhakal [18] reported higher risks of prematurity and neonatal asphyxia, attributed to untreated underlying conditions. Mascarello et al. [22] demonstrated an association with increased neonatal morbidity related to intrauterine scarring, while Keag et al. [24] reported increased risks of prematurity and low birth weight. Conversely, Oros Ruiz et al. [25] found that subsequent pregnancies could have favorable outcomes when adequately monitored, particularly in settings with strengthened antenatal care services.

## CONCLUSION

This study conducted at the Mono–Couffo Departmental Hospital Center highlighted a high prevalence of caesarean section, predominantly performed as emergency procedures. Although unfavorable maternal and fetal outcomes were relatively limited, they remain concerning due to their implications for maternal and neonatal health. Factors such as inadequate antenatal care, obstetric history, and certain maternal medical conditions were found to have a significant influence on these outcomes. These findings support the need to strengthen antenatal surveillance, improve planning of high-risk deliveries, and enhance the quality of obstetric care in order to reduce maternal and fetal morbidity in Benin.

## Data Availability

The data underlying the findings of this study contain sensitive patient information and cannot be made publicly available due to ethical and legal restrictions related to patient confidentiality under national regulations in Benin. Anonymized data may be made available upon reasonable request to the corresponding author and with permission from the Mono–Couffo Departmental Hospital Center and the relevant institutional authorities.

## ACKNOWLEDGEMENTS

The authors express their sincere gratitude to the Research Unit in Epidemiology and Population Health (CaRESaP; www.caresap.org) for its methodological, statistical, and logistical support throughout the conduct of this study. Their contribution was instrumental in ensuring the scientific rigor and quality of the analyses presented.

